# PROTEOMIC IDENTIFICATION OF NOVEL AUTOANTIBODY TARGETS IN RHEUMATOID ARTHRITIS AND DIAGNOSTIC PERFORMANCE OF TGF-β1-INDUCED PROTEIN AND SERUM AMYLOID A AUTOANTIBODIES

**DOI:** 10.64898/2026.09.25.26361099

**Authors:** Melissa Frenchmeyer, Michael Moleski, Allison Keilman, Stephanie Fernandez, Kilsun Kim, Keith Zientek, Phillip Wilmarth, Ashok Reddy, Lina Gao, Vishnupriya R. Paturi, Charles T. Roberts, Srinivasa R. Nagalla

## Abstract

**Objective:** The principal diagnostic test for rheumatoid arthritis (RA) relies on the detection of anti-cyclic citrullinated peptide (CCP) antibodies. The CCP test exhibits moderate sensitivity and specificity but does not adequately detect a high proportion of cases or effectively distinguish RA from related conditions such as systemic lupus erythematosus (SLE). To discover improved diagnostic biomarkers for RA, we performed a comprehensive proteomic analysis of antibody-bound antigens present in RA, SLE, and healthy control (HC) sera and identified proteins associated with transforming growth factor-β1 (TGF-β1) signaling and assessed the diagnostic performance of antibodies against these antigens in comparison to CCP.

**Methods:** Total antibodies were immunoprecipitated from pooled sera of HC individuals (n=3) and patients with clinically diagnosed RA (n=3) or SLE (n=3). Antibody-bound antigens were identified using quantitative bottom-up proteomics techniques. Antigens associated with TGF-β1 signaling were selected for evaluation, and corresponding serum autoantibodies were assessed for their ability to classify RA using a POC lateral-flow immunochromatographic format.

**Results:** Proteins present in antibody immunoprecipitations included TGF-β1-induced protein (TGFBI), thrombospondin 1 (TSP1), serum amyloid A1 (SAA), and fibronectin (FN). POC assays using these candidates, as well as TGF-β1 itself, as capture antigens were used to detect corresponding serum autoantibodies in 107 subjects with clinically diagnosed RA, 42 subjects with other rheumatic diseases (non-RA), and 121 HC subjects. Receiver operating characteristic curves were constructed using RA and HC groups to assess diagnostic performance, and optimal cutpoints identified using Youden’s Index (maximizing the sum of sensitivity and specificity). CCP test results were determined using previously established thresholds. Using an analysis set of 169 subjects in the HC and RA groups who had values for TGFBI, SAA, and CCP, TGFBI autoantibodies achieved a sensitivity of 82%, specificity of 98.8%, positive likelihood ratio (PLR) of 65.6, and NLR of 0.2. SAA autoantibodies achieved a sensitivity of 92.1%, specificity of 85%, PLR of 6.1, and NLR of 0.1. Both outperformed CCP (sensitivity of 68.5%, specificity of 95%, PLR of 13.7, and NLR of 0.3). Using an analysis set of 120 subjects in the RA and non-RA groups who had values for TGFBI, SAA, and CCP, TGFBI and SAA autoantibodies exhibited sensitivities and specificities of 82 and 92.1% and 74.2 and 45.2%, respectively, compared to 68.5 and 74.2% for CCP. Autoantibodies to TGF-β1, TSP1, and FN exhibited only modest diagnostic performance.

**Conclusion:** RA sera contain autoantibodies targeting multiple proteins associated with the TGF-β1 signaling pathway that is implicated in inflammatory processes in RA. Among the candidates tested, TGFBI and SAA autoantibodies exhibited the highest diagnostic performance in a POC lateral-flow format, supporting their potential utility as biomarkers for RA diagnosis. These findings also support further investigation of the TGF-β1 signaling pathway as a potential therapeutic target in RA.

## INTRODUCTION

Rheumatoid arthritis (RA) is a serious and debilitating autoimmune disease that elicits joint inflammation and attendant bone loss (1), although other organ systems are also affected (2). The global prevalence of RA is ∼0.5-1% (3,4), with women being predominately affected; however, the global burden of RA has been steadily increasing over recent decades (5). Other rheumatic diseases such as systemic lupus erythematosus (SLE) and Sjogren’s Syndrome (SS) can exhibit symptoms similar to those of RA, specifically fatigue, joint pain, and stiffness. While RA, SLE, and SS are all autoimmune diseases and share similar peripheral blood gene expression profiles (6), specific organ targets and therapeutic approaches vary. For example, RA requires disease-modifying antirheumatic drugs (DMARDs), SLE treatment involves suppression of systemic inflammation and autoimmune attacks, and SS treatment is limited to systemic immunosuppressants and local symptom management. Thus, correct diagnosis is crucial for the application of appropriate therapeutic approaches.

Diagnosis of RA relies on a combination of clinical assessment and biomarkers, principally rheumatoid factor (RF), comprised of autoantibodies directed against the Fc domain of IgG, and autoantibodies that recognize cyclic citrullinated peptides (CCP) (7) containing arginine residues that have been citrullinated. Current CCP tests employ cyclic citrullinated peptides derived from the sequence of filaggrin and selected other citrullinated proteins, and, while superior to RF alone, are also positive in related conditions such as SLE and SS (8). This has led to renewed interest in identifying additional biomarkers that can facilitate diagnosis, prognosis, assessment of disease severity, and response to therapy (9–17). Early diagnosis may also facilitate prevention through the identification of individuals without clinically apparent disease, but who may benefit from interventions to delay symptom onset (18).

We previously described the serendipitous identification of anti-citrullinated human serum albumin antibodies (ACA) in RA sera (19), confirming the previous report of Hefton et al. (20). Use of a prototype point-of-care (POC) test for ACA as a novel RA biomarker increased the rate of detection of RA over that obtained with CCP alone (19), findings replicated by Wang et al. using the same POC device (21). However, the combined sensitivity of ACA and CCP in our study was only 0.66 in that study population; thus, there exists significant room for further improvement in both sensitivity and specificity in RA biomarker testing. In this report, we describe our identification of components of the transforming growth factor (TGF)-β1 processing/signaling pathway in immune complexes from RA sera and the diagnostic performance of tests for autoantibodies directed against these antigens and the superior performance of anti-TGF-β1-induced protein (TGFBI) and serum amyloid A1 (SAA) autoantibodies in particular as novel biomarkers for RA diagnosis.

## METHODS

### Immunoprecipitation of serum antibody-antigen complexes

Immunoprecipitation of native immunoglobulins and their bound antigens in pooled sera from HC (n=3), RA (n=3), and SLE (n=3) subjects was performed using a 1:1 ratio of PureProteome Kappa and Lambda Ig Binder Magnetic Beads (Cat. #s LSKMAGKM02 and LSKMAGLM02, respectively, Millipore Sigma, Burlington, MA). For each sample, 150 µL each of kappa and lambda beads were combined and washed twice with 500 µL of IP wash buffer (1x PBS). Each pooled HC, RA, and SLE sample was diluted 1:4 in 1x PBS to a total volume of 100 µL and incubated with the pre-washed beads for 1 hour with end-over-end rotation at room temperature. The supernatant containing the unbound fraction was removed and the beads washed 3x with IP wash buffer. The bound antibody-antigen complexes were eluted using 175 µL of elution buffer (200 mM glycine-HCl, pH 2.6) with incubation for 10 minutes with end-over-end rotation. The eluted fractions were neutralized with 1/10 volume neutralizing buffer (2 M Tris, pH 8) and transferred on ice for analysis by mass spectrometry.

### Sample preparation

Sample eluates were dried, dissolved in 100 µL of buffer (5% sodium dodecyl sulfate and 50 mM triethyammonium bicarbonate (TEAB), pH 8), reduced with dithiothreitol at 37 °C for 15 min, alkylated with iodoacetamide for 15 min at room temperature, acidified with phosphoric acid followed by the addition of 90%/10% methanol/100 mM TEAB, pH 7.55. Samples were then loaded onto an S-trap micro column (Cat. #C002-MICRO, Protifi, Fairport, NY) and washed with 90%/10% methanol/100 mM TEAB, pH 7.55. Bound proteins were digested with 3.2 µg of trypsin (Cat. #90057, Thermo Fisher Scientific, Waltham, MA) at 37^0^C overnight. Peptides were eluted with 50 mM TEAB, 0.2% formic acid and 50%/50% acetonitrile/0.2% formic acid. Eluates were pooled and dried and dissolved in 50 µL of HPLC water. The peptide concentration was determined using a calorimetric assay kit (Cat. # 23227, Thermo Fisher Scientific).

### LC-MS/MS analysis

Five µL of each sample was diluted to 50 µL with 5% formic acid and 2.5 µL/sample were injected for reverse-phase C18 liquid chromatography. Chromatographic separation was performed using a Dionex UltiMate NCS-3500RS system (Thermo Scientific, San Jose, CA). Mobile phase A consisted of 0.1% formic acid in water and mobile phase B consisted of 0.1% formic acid in acetonitrile. Peptides were initially trapped on a Thermo Acclaim PepMap C18 trap column (100 µm x 2 cm, NanoViper, 5-µm particle size) and washed for 10 minutes with mobile phase A. Following the wash step, peptides were separated on a Thermo PepMap RSLC C18 analytical column (75 µm x 25 cm, EasySpray, 2-µm particle size). The flow rate delivered to the analytical column was maintained at 300 nL/min for a total run time of 90 min. A multi-step gradient elution profile was used to separate the peptides. From 0 to 5 min, the column was held at 2% mobile phase B to wash the samples on the trap column. At 5 min, the solvent composition was increased to 5% mobile phase B, followed by a linear gradient to 30% B over 70 min. The gradient was then increased to 98% mobile phase B for 5 min to wash the column. The mobile phase B composition was reduced back to 2% mobile phase B at 82 min, and the analytical column was equilibrated until the end of the run at 90 minutes.

Tandem mass spectrometry data were collected using an Orbitrap Eclipse Tribrid instrument (Thermo Scientific, San Jose, CA) configured with an EasySpray NanoSource. The nanospray ionization source was operated with a spray voltage of 2000 V, and the ion transfer tube temperature was maintained at 300 °C. Survey (MS1) scans were performed in the Orbitrap mass analyzer at a resolution of 60,000 with RunStartEasyIC mass calibration enabled. Survey scans from 350 to 1400 m/z were acquired in profile mode with a maximum injection time of 118 milliseconds and an automatic gain control (AGC) target of 4×10^5^ ions. Dynamic exclusion was enabled for 45 sec with 10 ppm tolerance, and charge states from 2 to 7 were included for fragmentation. Isotope exclusion was enabled and peptide matching was enabled. MS/MS (MS2) scans were also acquired in the Orbitrap analyzer. Precursor ions exceeding the intensity threshold of 5000 were selected using a 1.6 m/z quadrupole isolation window with no offset, followed by higher-energy collisional dissociation (HCD) fragmentation at a normalized collision energy of 30%. Centroided MS2 spectra were acquired at a resolution of 7,500 with a maximum injection time of 50 milliseconds, an AGC target of 5×10^4^ ions, and using auto scan range. MS1 and MS2 cycle time was 2 sec.

### MS Data Analysis

The binary instrument files were processed with the PAW pipeline (22). Binary files were converted to text files using MSConvert (23). Python scripts fragment ion spectra in MS2 format (24). The Comet search engine (version 2016.03) (25) was used: 1.25-Da monoisotopic peptide mass tolerance, 0.02-Da monoisotopic fragment ion tolerance (bin offset of 0.0), fully tryptic cleavage with up to three missed cleavages, variable oxidation of methionine residues (+15.9949 Da), and static alkylation of cysteines (+57.0245 Da). Searches used UniProt proteome UP000005640 (human, taxon ID 9606) canonical FASTA sequences (20,598 proteins) downloaded in April 2024. Common contaminants (175 sequences excluding any albumins) were added, and sequence-reversed entries were concatenated for a final protein FASTA file of 41,546 sequences.

Top-scoring peptide spectrum matches (PSMs) were filtered to a 1% false discovery rate (FDR) using interactive delta-mass and conditional Peptide-prophet-like linear discriminant function scores (26). Incorrect delta-mass and score histogram distributions were estimated using the target/decoy method (27). The filtered PSMs were assembled into protein lists using basic and extended parsimony principles and required two distinct peptides per protein. The final list of identified proteins, protein groups, and protein families were used to define unique and shared peptides for quantitative use. Corrected protein spectral counts (SpC) were computed where shared peptide counts were fractionally split based on relative unique peptide counts. The corrected SpC values were used for comparisons between samples.

Data analysis was performed using Proteome Discoverer 3.1 using Sequest (v. 2016.01, rev. 3) against the September 2024 download of the canonical homo sapiens Uniprot FASTA sequences (one sequence per gene) and concatenated sequence-reversed entries to estimate error thresholds (1% FDR) and 179 common contaminant sequences and their reversed forms.

### Autoantibody lateral-flow POC test format

Serum or plasma samples were analyzed using a POC test system (ModalityRx, Inc., Hillsboro, OR). The test system consists of an immunochromatography test strip in a cassette and a portable reader for quantification. Test strips were configured with antibodies against human TGF-β1 (Cat. # MA1-169, Thermo Fisher Scientific), TGFBI (Cat. # AF2935, R&D Biosystems, Minneapolis, MN), thrombospondin 1 (TSP-1) (Cat. # MA5-13398, Thermo Fisher Scientific), SAA (Cat. # 100806, Medix Biochemica, St. Louis, MO), or fibronectin (FN) (ModalityDx, Hillsboro, OR) bound to a nitrocellulose membrane. biomarker-specific immune complexes in serum samples were detected with a colloidal gold anti-human IgA/G/M cocktail (**Figure 1**). 150 µL of 1:40 diluted serum or plasma was added to the test strip and inserted into a proprietary reader that utilizes image analyses for quantification. The concentration is displayed at the end of 15 minutes. Results for TGFBI and SAA autoantibodies are reported as arbitrary U/mL, with values >54 for TGFBI autoantibodies and >30 for SAA autoantibodies considered positive using the cutpoints determined as described below. The dynamic range of the assay is 15-4,000 U/mL, and intra- and inter-assay coefficients of variation are 5.1 and 8.7%, respectively.

**Figure 1.**
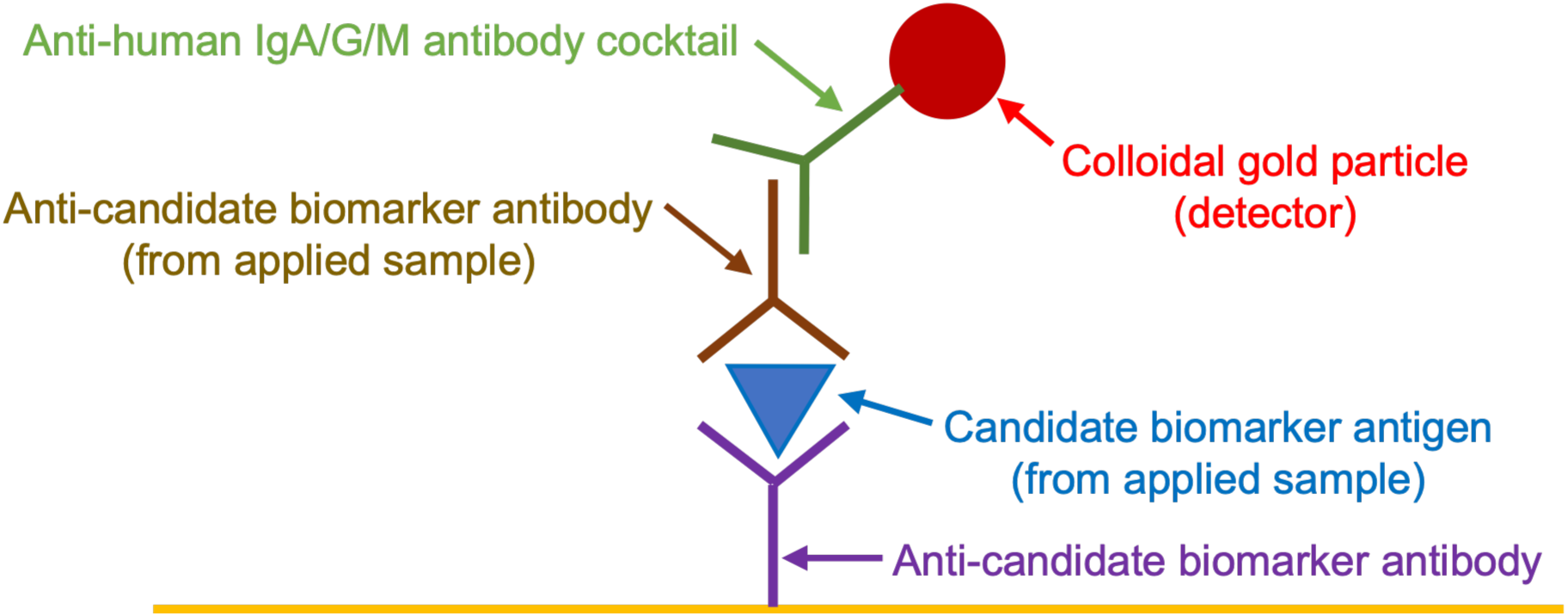
**Schematic of POC test strip architecture for performance assessment of novel RA biomarkers.**

### Determination of sensitivity and specificity of RA-enriched autoantibody antigen tests for RA diagnosis vs CCP

The prospective study population employed in this study overlapped with that used in our previous analysis of ACA (29) and was comprised of 107 subjects with clinically diagnosed RA, 42 subjects with other rheumatic disease (non-RA; 16 SLE, 2 RA/SLE, 1 spondyloarthritis, 1 gout, 1 psoriatic arthritis, 1 ankylosing spondylitis, 1 polyarthritis, 11 Graves’ disease, and 8 Hashimoto’s disease), and 39 healthy control (HC) samples from an East Asian population. An additional 80 HC samples from a US population were obtained from Innovative Research, Inc. (Novi, MI) for a total of 119 total HC samples. General characteristics of the HC, RA, and non-RA groups are shown in **Table 1**. The number of subjects tested in **Figures 3 and 4** was slightly less than the total in each category due to insufficient sample volumes available for all assays.

**Figure 2.**
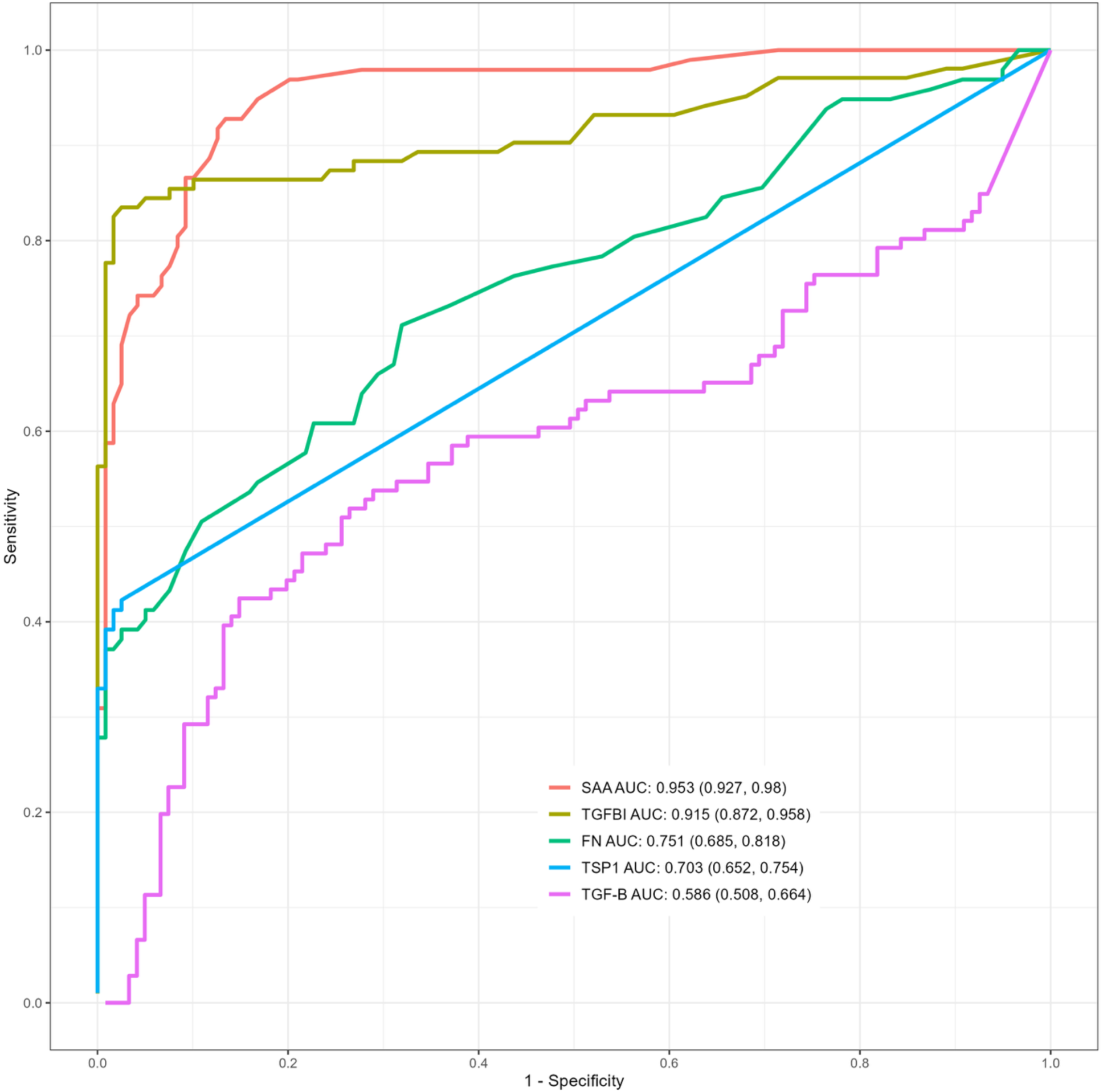
**Comparison ROC curves of candidate TGF-**β**1 pathway autoantibody biomarkers.** Performance measures are shown with 95% confidence intervals.

**Figure 3.**
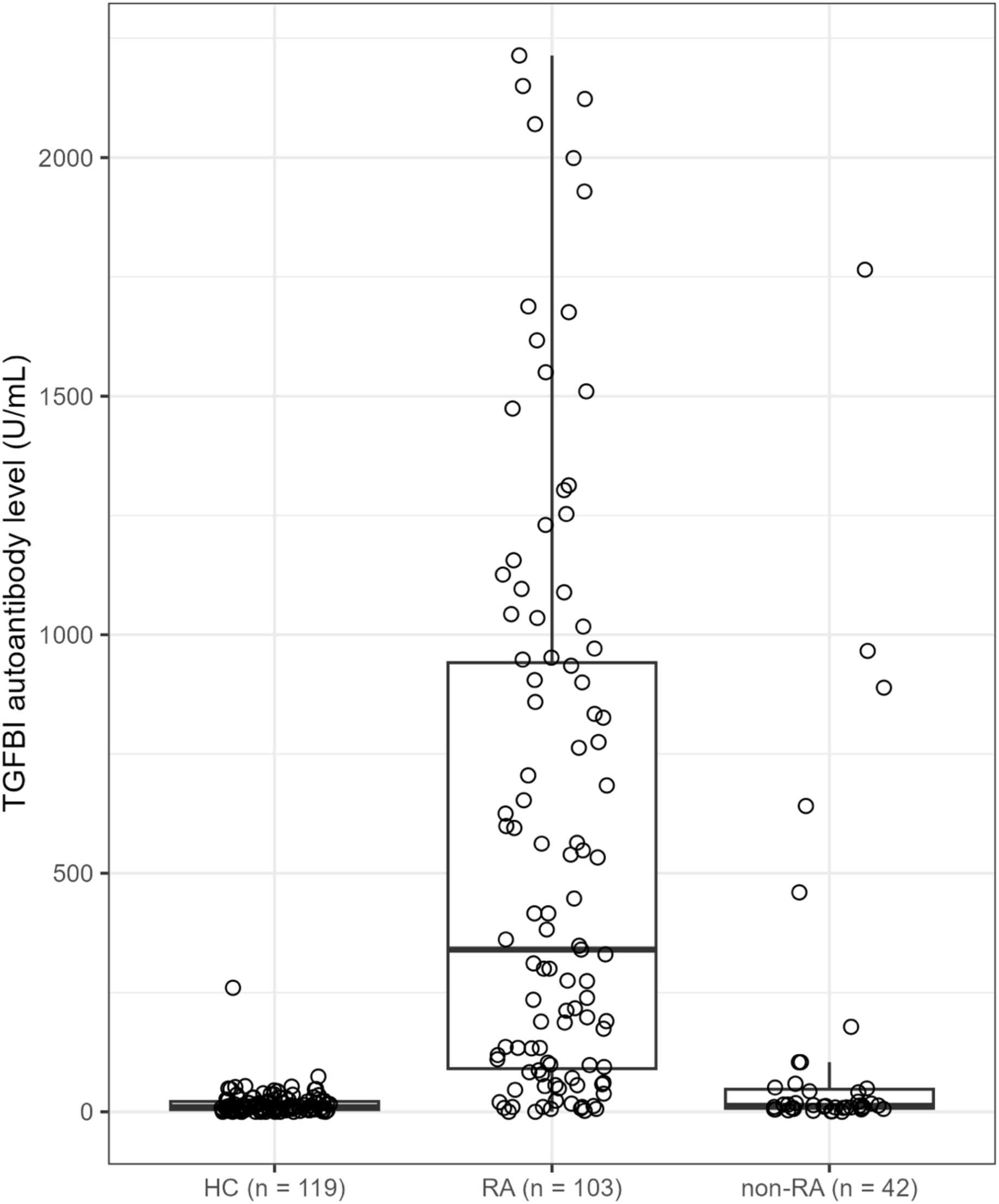
TGFBI autoantibody levels in all participants. Kruskal-Wallis test p-values were <0.0001 in pairwise comparisons of HC and RA and RA and non-RA, but 0.25 for HC and non-RA (Wilcoxon rank sum test followed by Bonferroni adjustment).

**Figure 4.**
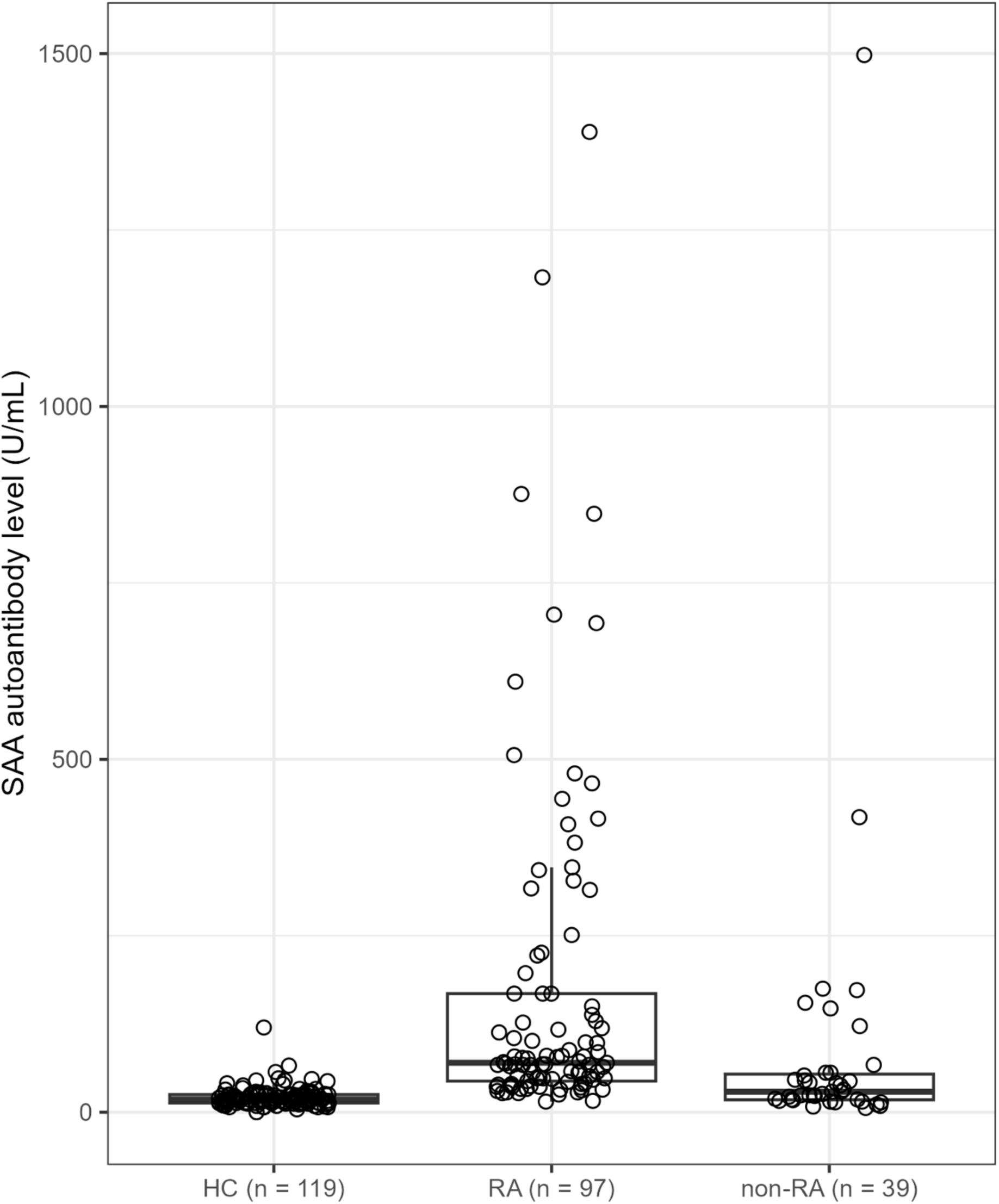
SAA autoantibody levels in all participants. Kruskal-Wallis test p-values were <0.0001 in pairwise comparisons of HC and RA and RA and non-RA, and 0.0008 for HC and non-RA (Wilcoxon rank sum test followed by Bonferroni adjustment).

**Table 1.** Participant characteristics.

| **Variable** | **HC** | **RA** | **Non-RA** | **p-value** |
| --- | --- | --- | --- | --- |
| N | 119 | 107 | 42 |  |
| Age (mean (SD)) | 38.64 | 48.63 | 35.60 | <0.001 |
| Sex = M (%) | 55 (45.5) | 10 (9.2) | 9 (21.4) | <0.001 |

Similarly, 169 samples in the HC (n=80) and RA (n=89) groups and 120 subjects in the RA (n=89) and RA (n=31) groups that had values for TGFBI and SAA autoantibodies and CCP were used to determine relative diagnostic performance as shown in **Table 3**. Samples were tested with the POC device described above as well as with a commercial CCP assay (Siemens Healthcare Diagnostics, Tarrytown, NY; positive value is >5 U/ml). Institutional IRB approval was obtained (CTRI/2020/10/028596, Kumudini Devi Diabetes Research Center Ramdevrao Hospital, Kukatpally, Hyderabad), the study was registered with appropriate regulatory authorities, and informed consent was obtained from all study participants.

### Statistical analyses

Receiver operating characteristic (ROC) curves were constructed to assess diagnostic performance of each biomarker, and the area under the ROC curve (AUC) are reported with 95% confidence intervals. Optimal cutpoints were identified using Youden’s Index (28). Prediction performance measures at the optimal cutpoints are reported with 95% exact Clopper-Pearson confidence intervals (29).

## RESULTS

### Autoantibody antigens present in RA, SLE, and HC sera

Quantitative bottom-up proteomic analysis of total IgG, M, and A immunoprecipitates from RA, SLE, and HC sera was used to detect novel autoantigens. **Table 2** shown the non-Ig antigens that were present in HC, RA, and SLE serum immune complexes. Among these were factors that modulate the TGF-β1 signaling pathway, including TGFBI, SAA, TSP1, and FN, each of which has been previously associated with RA as well as being directly or indirectly involved in TGF-β1 action as described below. Accordingly, we focused on autoantibodies directed against these proteins as well as TGF-β1 itself in the assessment of diagnostic performance detailed below.

**Table 2.**
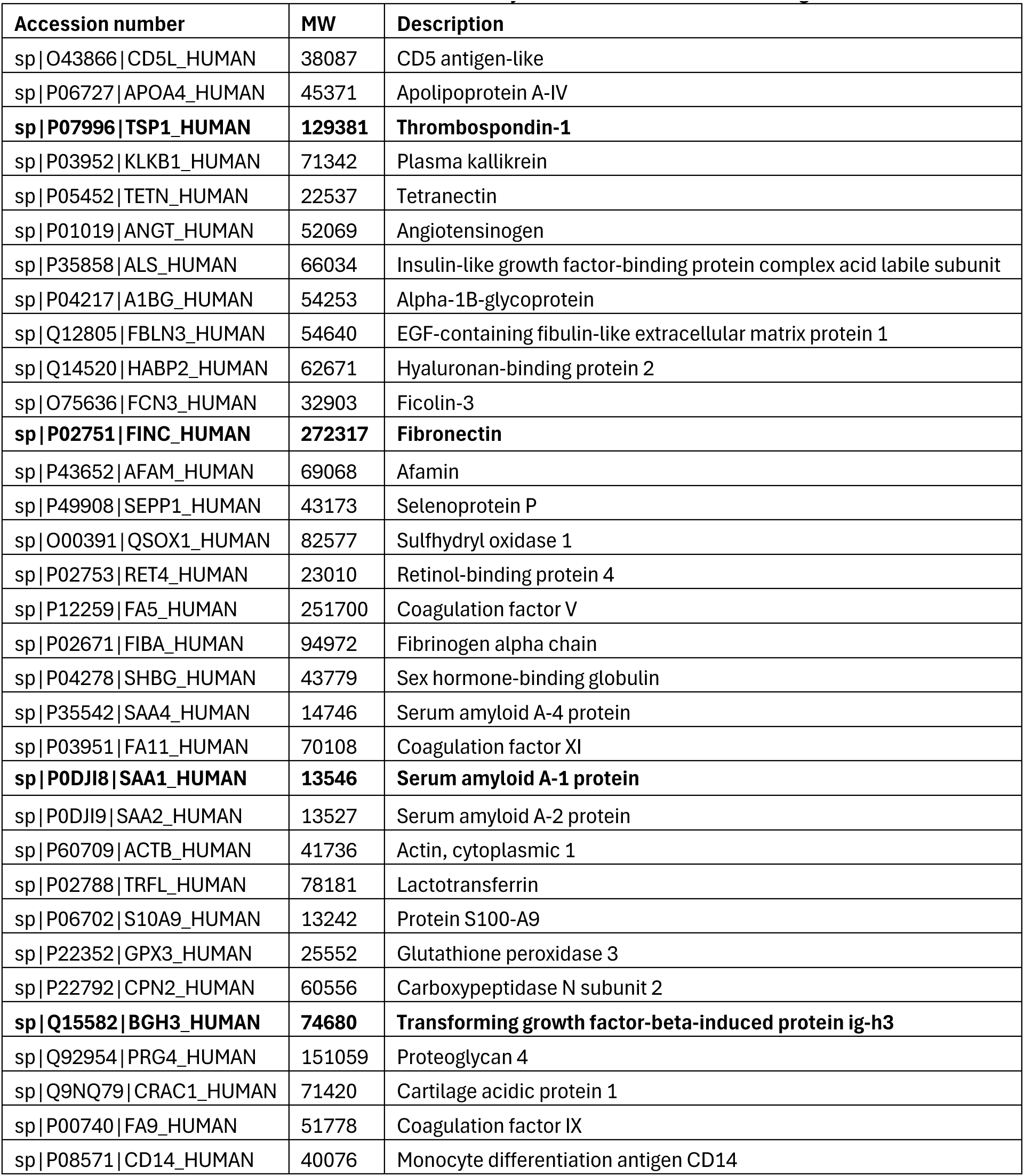
Antigens present in immune complexes isolated from HC, RA, and SLE serum pools. Immunoglobulins excluded from this list. TGF-β1 pathway-related autoantigens shown in bold were selected for assessment of their utility as biomarkers for RA diagnosis.

**Table 3.**
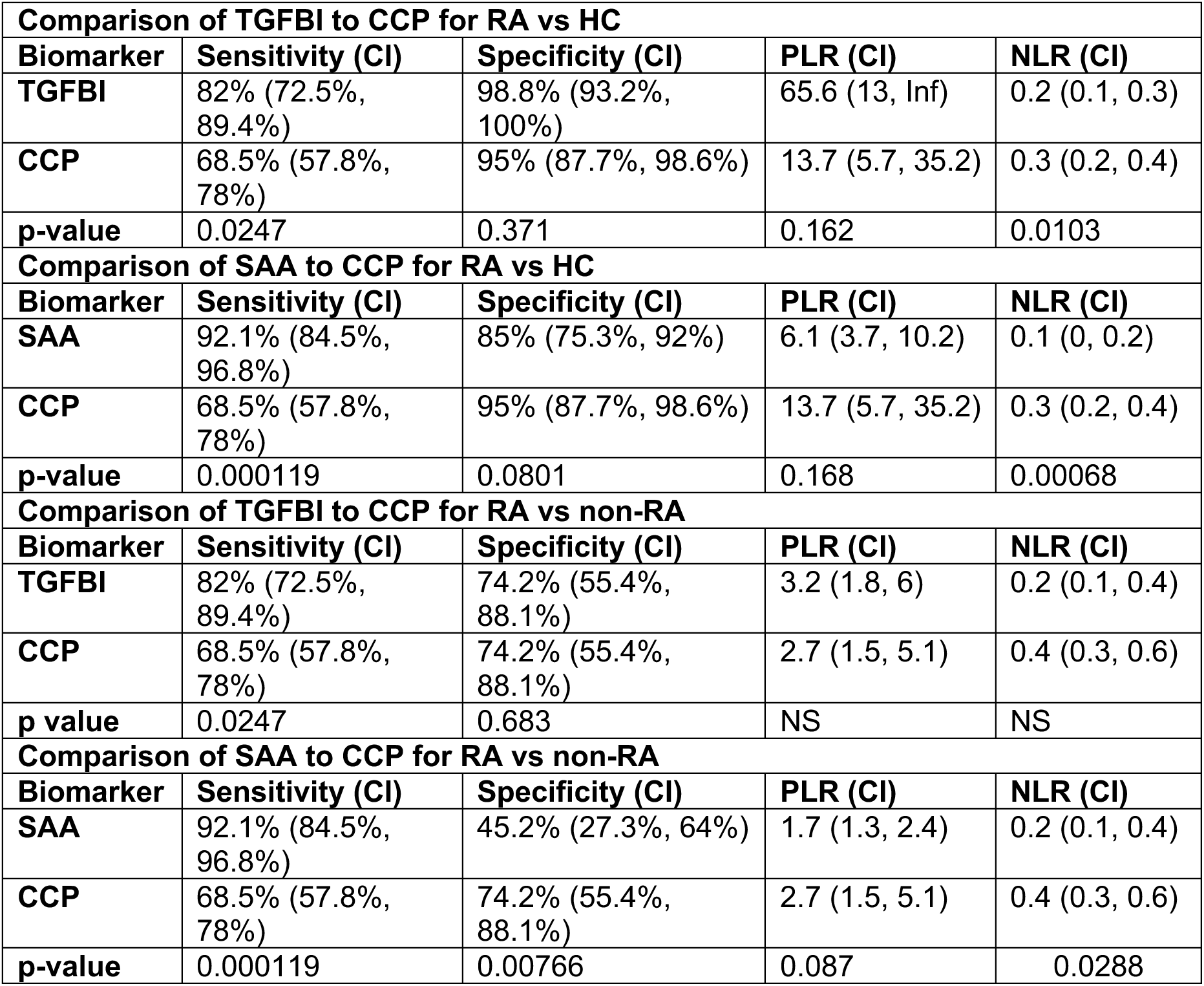
Diagnostic performance parameters of TGFBI autoantibodies vs CCP. Exact binomial confidence limits (95% CI) are shown. Comparisons between markers were performed using McNemar’s test with continuity correction. NS, not significant.

### Diagnostic performance of novel RA biomarkers

Receiver operating characteristic (ROC) curves were constructed using RA and HC groups to assess diagnostic performance, and optimal cutpoints identified using Youden’s Index (maximizing the sum of sensitivity and specificity). Based on the ROC curves shown in **Figure 2**, TGFBI and SAA autoantibodies exhibited superior performance to those directed against TGF-β1, TSP1, and FN. Specifically, TGFBI and SAA autoantibody AUCs were 0.915 and 0.953, respectively, while corresponding AUCs for FN, TSP1, and TGF-β1 were 0.751, 0.703, and 0.586, respectively. Thus, autoantibodies to TGF-β1, TSP1, and FN exhibited only modest diagnostic performance.

As shown in **Figure 3**, median TGFBI autoantibody levels in HC, RA, and non-RA sera were 10.0, 340.0, and 12.5 U/mL; thus, the median level in RA sera was 34-fold higher than in HC sera (p<0.0001), 27-fold higher than in non-RA sera (p<0.0001), and 6.3-fold higher than the optimal cutpoint of 64.1 U/mL calculated as described above. Median TGFBI autoantibody levels in HC and non-RA sera were not statistically different (p=0.25). **Supplemental Figure 1** shows the average TGFBI autoantibody levels in female and male participants in the RA and non-RA groups. TGFBI autoantibody levels were significantly lower in the male than in the female participants in the RA group (p=0.00762), while levels in the non-RA group were lower than in the RA group as also shown in **Figure 3** but were not different between females and males (p=0.372).

As shown in **Figure 4**, median SAA autoantibody levels in HC, RA, and non-RA sera were 18, 70, and 29 U/mL, respectively; thus, the median level of SAA autoantibodies in RA sera was 3.8-fold higher than in HC sera (p<0.0001), 2.4-fold higher than in non-RA sera (p<0.0008), and 2.3-fold higher than the calculated optimal cutpoint of 30 U/mL. In contrast to TGFBI autoantibodies, the median level of SAA autoantibodies in non-RA sera was statistically greater than the level in HC sera (p<0.0001). **Supplemental Figure 2** shows the average SAA autoantibody levels in female and male participants in the RA and non-RA groups. SAA autoantibody levels were significantly lower in the male than in the female participants in the RA group (p=0.0235), while levels in the non-RA group were lower than in the RA group as also shown in **Figure 4** but were not different between females and males (p=0.289).

While TGFBI and SAA autoantibodies were both characterized by a high AUC in the ROC curves shown in **Figure 2** (0.915 for TGFBI autoantibodies and 0.953 for SAA autoantibodies), median TGFBI autoantibody levels were higher than median SAA autoantibody levels in RA sera.

### Novel biomarker performance in comparison to CCP

**Table 3** lists the sensitivity, specificity, positive likelihood ratios ((PLR), and negative likelihood ratios (NLR) for the TGFBI and SAA autoantibodies obtained using the POC test as well as the same parameters determined using a commercial assay for CCP and previously established thresholds. In distinguishing RA from HC samples, TGFBI autoantibodies achieved a sensitivity of 82%, specificity of 98.8%, PLR of 65.6, and NLR of 0.2 at the optimal cutpoint. SAA autoantibodies achieved a sensitivity of 92.1%, specificity of 85%, PLR of 6.1, and NLR of 0.1 at the optimal cutpoint. Both outperformed CCP overall (sensitivity of 68.5%, specificity of 95%, PLR of 13.7, and NLR of 0.3).

TGFBI and SAA autoantibodies were also superior to CCP in distinguishing RA from non-RA samples. Specifically, TGFBI autoantibodies achieved a sensitivity of 82%, specificity of 74.2%, PLR of 3.2, and NLR of 0.2 at the optimal cutpoint. SAA autoantibodies achieved a sensitivity of 92.1%, specificity of 45.2%, PLR of 1.7, and NLR of 0.2 at the optimal cutpoint. Both outperformed CCP overall (sensitivity of 68.5%, specificity of 74.2%, PLR of 2.7, and NLR of 0.4).

We also analyzed the relationship between the presence of TGFBI and SAA autoantibodies and those recognized by the commercial CCP test. Of the 107 RA sera, 95 had sufficient sample volume to be tested for both TGFBI autoantibodies and CCP. As shown in **Figure 5**, Of the 95 RA sera tested, 85 (89%) were positive for either TGFBI autoantibodies or for CCP. Of these, 78 (91%) were TGFBI-positive and 66 were CCP-positive (78%). Importantly, 19 RA sera were TGFI-positive but CCP-negative, while only 7 RA sera were CCP-positive but TGFBI-negative. Thus, 24% of RA samples in which TGFBI autoantibodies were present were CCP-negative.

**Figure 5.**
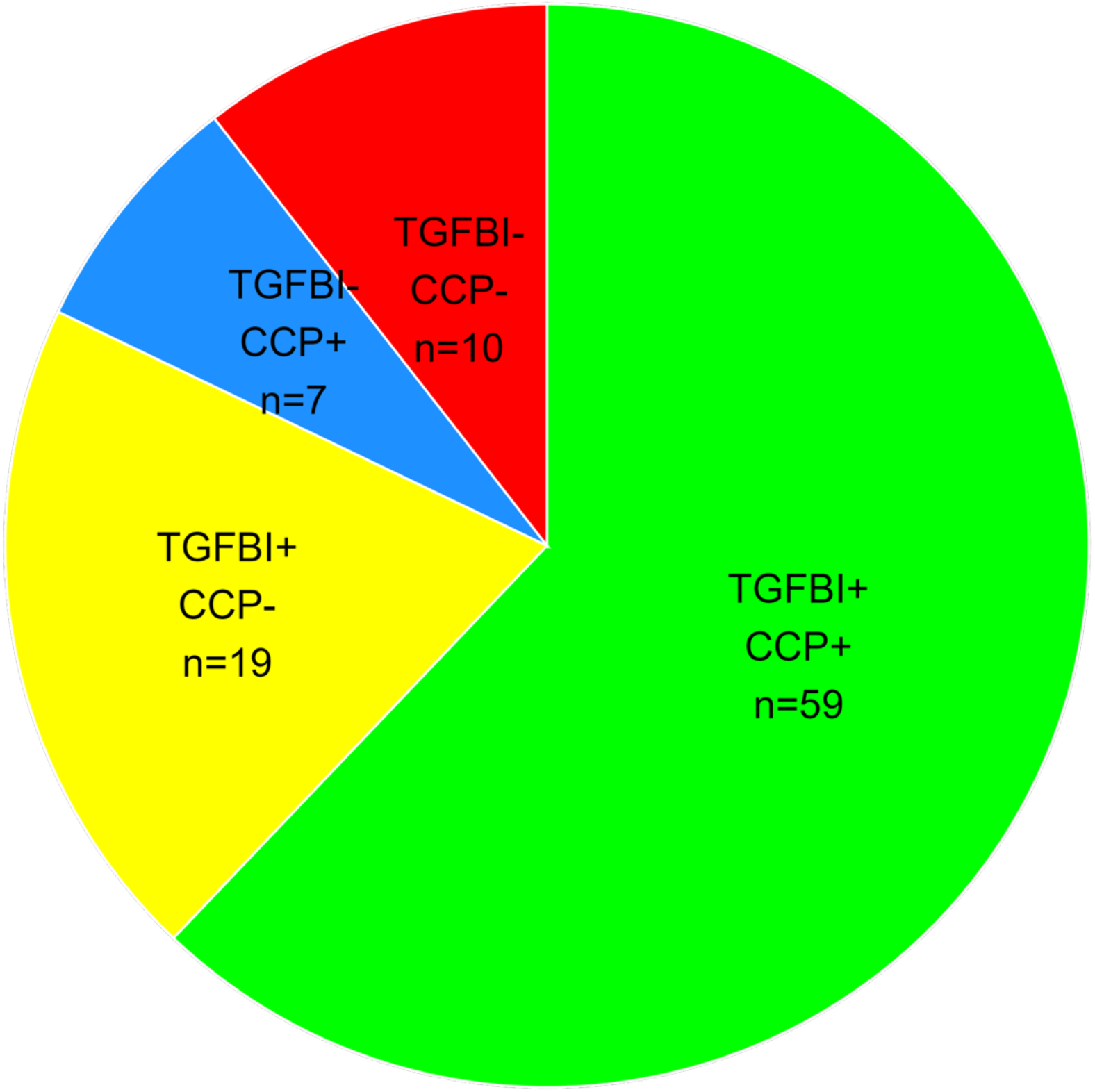
TGFBI autoantibody specificity and relationship to CCP. Pie chart depicts RA subpopulations segregated by TGFBI and CCP status.

A similar assessment of SAA autoantibodies is shown in **Figure 6**. Of the 107 RA sera, 89 had sufficient sample volume to be tested for both SAA autoantibodies and CCP. As shown in **Figure 6**, of the 89 RA sera tested, 84 (94%) were positive for either SAA autoantibodies or for CCP. Of these, 81 (91%) were SAA-positive and 61 were CCP-positive (68%). Additionally, 23 RA sera were SAA-positive but CCP-negative, while only 3 RA sera were CCP-positive but SAA-negative. Thus, 26% of RA samples in which SAA autoantibodies were present were CCP-negative.

**Figure 6.**
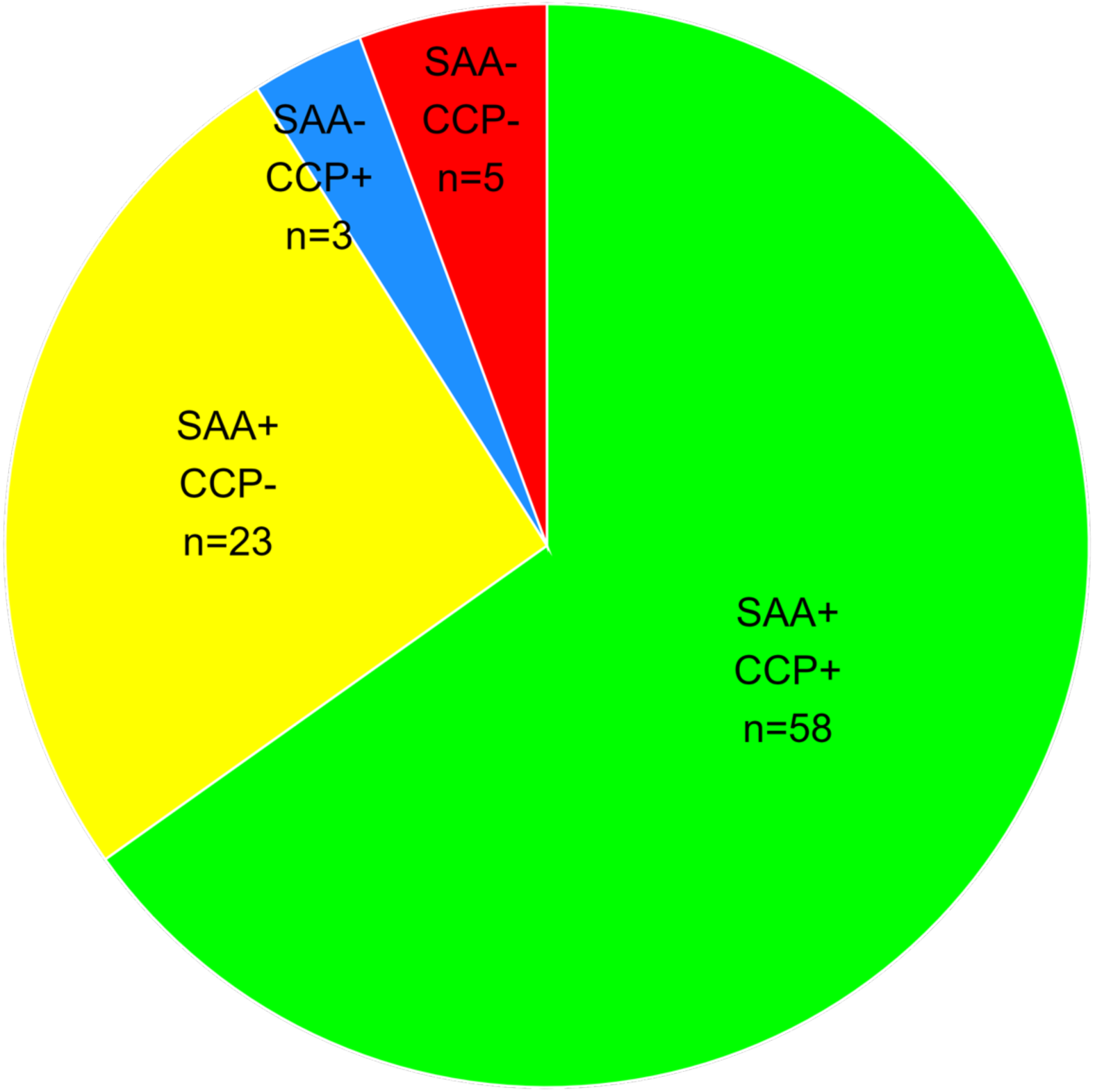
SAA autoantibody specificity and relationship to CCP. Pie chart depicts RA subpopulations segregated by SAA and CCP status.

These data demonstrate that both TGFBI and SAA autoantibodies extend the proportion of RA samples diagnosed by CCP alone and exhibit greater diagnostic performance than CCP as standalone diagnostic biomarkers.

## DISCUSSION

There is a continuing need for improved biomarkers for RA diagnosis to complement clinical characterization of patient symptoms and for potential prediction of response to therapy. Recent RA therapeutic approaches include biologic DMARDS for inhibition of tumor necrosis factor and interleukin-6 pathways and targeted synthetic DMARDS such as tofacitinib, baricitinib, and upadacitinib for inhibition of the Janus kinase pathway (30–32), with mesenchymal stem cell therapies under development (33–36). Given the array of current and emerging therapeutic approaches, a variety of new biomarkers may be required.

RA pathogenesis is thought to involve multiple signaling pathways (37), and previous biomarker discovery has employed proteomic approaches as described above (10,15,16) as well as gene expression (38), and metabolomic (39) profiling of patient samples. In this study, we focused specifically on the circulating antigens targeted by autoantibodies present in RA, SLE, and HC sera to identify novel biomarkers amenable to POC testing and to reveal additional pathways that may contribute to RA initiation, progression, or severity. The rationale for assessment of circulating biomarkers is based on the practical utility of plasma biomarkers for diagnostic testing and the similarity between the plasma and synovial proteomes in RA based on the hyperpermeability of the RA synovial compartment (15).

Early data on the potential role of TGF-β1 in RA were mixed and based principally on animal and in vitro studies (40–42), but recent data implicate TGF-β1 in RA-associated pulmonary fibrosis (43) and treatment resistance (44). TGFBI is a downstream effector of TGF-β1 action (45) that is upregulated in RA synovial tissue (46). TSP-1 modulates TGF-β1 processing (47,48) and is upregulated in RA synovial tissue and plasma and implicated in RA pathogenesis (49–54). SAA is also involved in TGF-β1 processing (55,56) and is a reported RA biomarker (57–59). FN modulates TGF-β1 processing through its interaction with latent TGF-β1 binding protein (60) and citrullinated FN autoantibodies have been described in RA (61). Thus, all the analytes focused on in our study directly or indirectly affect TGF-β1 action and are each implicated in RA. These data suggest that TGF-β1 action has a major role in RA pathogenesis and that the TGF-β1 pathway may represent an under-addressed therapeutic target as previously suggested by Rico et al. (50) and recently explored by Aarts et al. (62).

Our comparison of the performance of the TGF-β1 pathway-associated autoantibodies revealed by our quantitative bottom-up proteomic analysis with that of CCP showed that, despite their presence in RA sera, autoantibodies directed against TGF-β1 itself, TSP-1, and FN were not useful as diagnostic biomarkers. On the other hand, TGFBI autoantibodies exhibited sensitivity, specificity, PLR and NLR values superior to CCP and SAA autoantibodies exhibited superior sensitivity and NLR values in comparison to CCP in this study population.

The differences in performance for the analytes investigated here, principally TGFBI and SAA autoantibodies and CCP, should be considered preliminary due to the small overall sample size. Our conclusions should be confirmed by future analyses of larger sample sets. Additionally, since the studies reported here principally employed a South Asian population in which 90% of the RA participants were female, it will be important to evaluate the diagnostic performance of TGFBI autoantibodies in additional patient populations with a higher representation of male participants, even though the prevalence of RA is higher in females (3,4). A separate limitation of our study is the lack of detailed information on the clinical course of the disease and symptomology of the RA cohort tested.

Although our findings support the utility of TGFBI and SAA autoantibodies as novel and efficacious biomarker for RA diagnosis with high sensitivity and specificity, further studies should determine whether these biomarkers can predict response to existing RA therapies as well as emerging mesenchymal stem cell approaches. In this context, Jiang et al. (63) have recently employed metabolomic analysis to characterize potential biomarkers of response to mesenchymal stem cell transplantation in SLE.

## Data Availability

All data produced in the present study are available upon reasonable request to the authors.

## ACKNOWLEDGEMENTS

Mass spectrometric analysis was performed by the OHSU Proteomics Shared Resource (RRID: SCR_009991) with partial support from NIH grants P30EY010572, P30CA069533, and S10 OD028533.

## ABBREVIATIONS

ACA: anti-citrullinated human serum albumin antibodies
CCP: cyclic citrullinated peptides
DMARDS: disease-modifying antirheumatic drugs
LR: likelihood ratio
LC-MS/MS: liquid chromatography/tandem mass spectroscopy
POC: point-of-care
RA: rheumatoid arthritis
ROC: receiver operating characteristic
SAA: serum amyloid protein 1
SLE: systemic lupus erythematosus
SS: Sjogren’s Syndrome
TGF-1: transforming growth factor 1
TGFBI: TGF-1-induced protein
TSP1: thrombospondin 1

---

**Supplemental figure 1.**
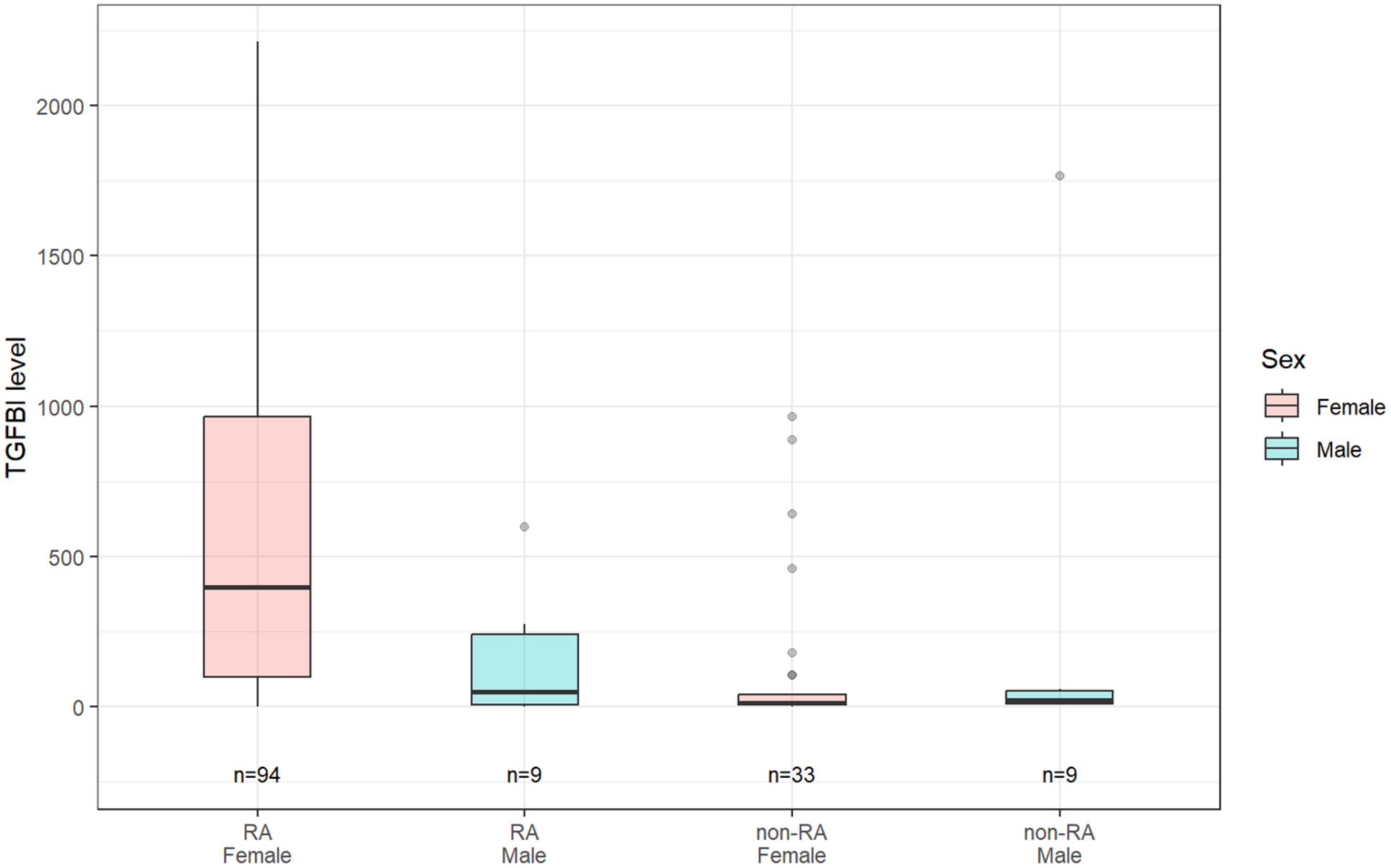
TGFBI autoantibody levels in female and male participants in the RA and non-RA groups. P values for pairwise comparisons of female and male participants in the RA and non-RA groups were 0.00762 and 0.374, respectively (Wilcoxon rank sum test).

**Supplemental figure 2.**
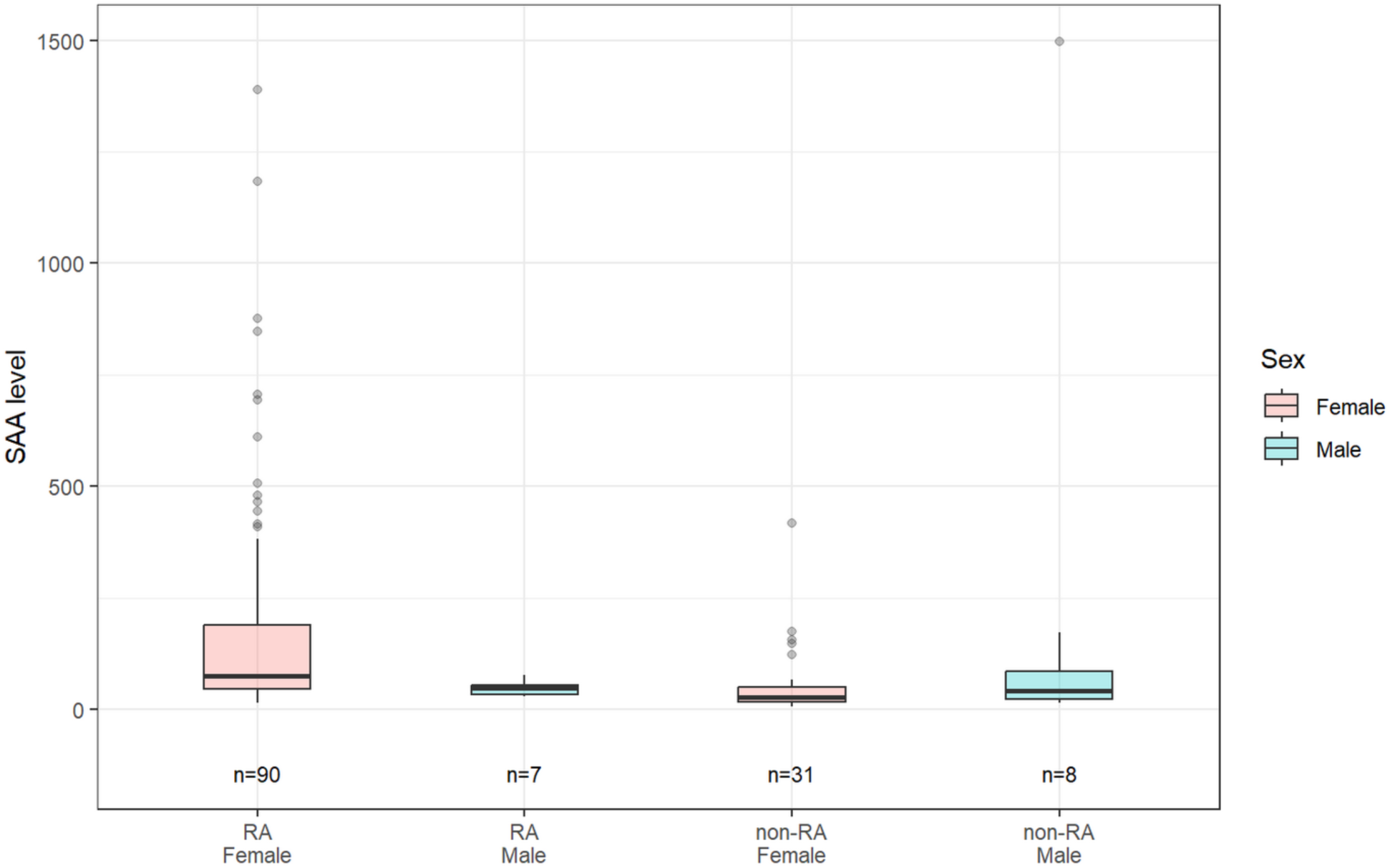
SAA autoantibody levels in female and male participants in the RA and non-RA groups. P values for pairwise comparisons of female and male participants in the RA and non-RA groups were 0.0235 and 0.289, respectively (Wilcoxon rank sum test).

## REFERENCES

1. Scott DL, Wolfe F, Huizinga TW. Rheumatoid arthritis. Lancet 2010;376:1094–1108.

2. Conforti A, Di Cola I, Pavlych V, et al. Beyond the joints, the extra-articular manifestations in rheumatoid arthritis. Autoimmun Rev 2021;20:102735.

3. Pedersen JL, Kjaer NK, Svendsen AJ, et al. Incidence of rheumatoid arthritis from 95 to 2001: Impact of ascertainment from multiple sources. Rheumatol Int 2009;29:411–415.

4. Collaborators, G.B.D.R.A., Global, regional, and national burden of rheumatoid arthritis, 1990–2020, and projections to 2050: A systematic analysis of the Global Burden of Disease Study 2021. Lancet Rheumatol 2023;5:e594–e610.

5. Uke P, Maharaj A, Adebajo A. A review on the epidemiology of rheumatoid arthritis: An update and trends from current literature. Best Prac Res Clin Rheumatol 2025;39:102036.2025

6. Toro-Dominguez D, Carmona-Saez P, Alarcon-Riquelme ME. Shared signatures between rheumatoid arthritis, systemic lupus erythematosus and Sjögren’s syndrome uncovered through gene expression meta-analysis. Arthritis Res Ther 2014;16:489.

7. Ronnelid J, Turesson C, Kastborn A. Autoantibodies in rheumatoid arthritis-laboratory and clinical perspectives. Front Immunol 2021;12:685312.

8. Payet J, Goulvestre C, Biale L, et al. Anticyclic citrullinated peptide antibodies in rheumatoid and nonrheumatoid rheumatic disorders: experience with 1162 patients. J Rheumatol 2014;41:2395–2402.

9. Shapiro SS. Biomarkers in rheumatoid arthritis. Cureus 2021;13:e15063.

10. Hu C, Dai Z, Xu J et al. Proteome profiling identifies serum biomarkers in rheumatoid arthritis. Front Immunol 2022;13:865425.

11. Pascual-Garcia S, Martinez-Peinado P, Lopez-Jaen AB et al. Analysis of novel immunological biomarkers related to rheumatoid arthritis disease severity. Int J Molec Sci 2023;24:12351.

12. Sahin D, Di Matteo A, Emery P. Biomarkers in the diagnosis, prognosis and management of rheumatoid arthritis: A comprehensive review. 2025;6:3–21.

13. Xue M, Wang H, Campos F, Jackson CJ, March L. Rheumatoid arthritis: biomarkers and the latest breakthroughs. Int J Molec Sci 2025;26:10594.

14. Neto M, Mendes B, Albuquerque F, da Silva JAP. Novel biomarkers in RA: Implication for diagnosis, prognosis, and personalised treatment. Best Prac Res Clin Rheumatol 2025;39:102021.

15. Stork EM, Nteak SK, van Rijswijck DMH et al. Multitiered proteome analysis displays the hyperpermeability of the rheumatoid synovial compartment for plasma proteins. Mol Cell Proteomics 2025;24:100900.

16. He S, Zhu C, Liu Y et al. A longitudinal cohort study uncovers plasma protein biomarkers predating clinical onset and treatment response of rheumatoid arthritis. Nature Comm 2025;16:6692.

17. Szekanecz Z, Gunkl-Toth L, Szamosi S et al. Diagnostic, prognostic and therapeutic biomarkers in rheumatoid arthritis. Nat Rev Rheumatol 2026;22:496–513.

18. Haville S, Deane KD. Pre-RA: Can early diagnosis lead to prevention? Best Clin Prac Res. Rheumatol 2021;36:101737.

19. Paturi VR, Uppuluri RR, Gao L, Roberts CT, Nagalla SR. Anti-citrullinated albumin antibodies as biomarker for the diagnosis of rheumatoid arthritis. Rheumatol: Curr Res 2024;14:362.

20. Hefton A, Kiang SY, Ni K, et al. Autoantibodies against citrullinated serum albumin in patients with rheumatoid arthritis. J Trans Autoimmun 2019;2:100023.

21. Wang X, Laine A, Chung S, Mustelin T. Diagnostic and prognostic utility of anti-modified albumin autoantibodies in rheumatoid arthritis. Arch Rheum Arth Res. 2022;2:000535.

22. Wilmarth PA, Riviere MA, and David LL. Techniques for accurate protein identification in shotgun proteomic studies of human, mouse, bovine, and chicken lenses. J Ocul Biol Dis Informat 2009;2:223–234.

23. Chambers MC, Maclean B, Burke R et al. 2012. A cross-platform toolkit for mass spectrometry and proteomics. Nature Biotech 2012;30:918–920.

24. McDonald WH, Tabb DL, Sadygov RG et al. 2004. MS1, MS2, and SQT-three unified, compact, and easily parsed file formats for the storage of shotgun proteomic spectra and identifications. Rapid Comm Mass Spect 2004;18:2162–2168.

25. Eng JK, Jahan TA, Hoopmann MR. 2013. Comet: an open-source MS/MS sequence database search tool. Proteomics 2013;13:2–24.

26. Keller A, Nesvizhskii AI, Kolker E, Aebersold R. Empirical statistical model to estimate the accuracy of peptide identifications made by MS/MS and database search. Anal chem 2002;74:5383–5392.

27. Elias JE Gygi SP. Target-decoy search strategy for increased confidence in large-scale protein identifications by mass spectrometry. Nat Meth 2007;4:207–214.

28. Youden WJ. Index for rating diagnostic tests. Cancer 1950;3:32–35.

29. Clopper CJ, Pearson ES. The use of confidence or fiducial limits illustrated in the case of the binomial. Biometrics 1934;26:404–413.

30. Domanska-Poboza J, Wislowska M. Evolving strategies in the treatment of rheumatoid arthritis: a historical perspective. Rheumatologia 2025;63:116–130.

31. Brown P, Pratt AG, Hyrich KL. Therapeutic advances in rheumatoid arthritis. Brit Med J 2024;384: e070856.

32. Nascimento MMP, Azuelos A, Nobrega ILP et al. State of the art in novel treatment strategies in rheumatoid arthritis: a brief review. Mediterr J Rheumatol 2025;36:149–158.

33. Mesa LE, Lopez JG, Quiceno LL et al. Safety and efficacy of mesenchymal stem cells therapy in the treatment of rheumatoid arthritis disease: A systematic review and meta-analysis of clinical trials. PLOS One 2023;18:e0284828.

34. Shimizu Y, Ntege EH, Azuma C et al. Management of rheumatoid arthritis: possibilities and challenges of mesenchymal stromal/stem cell-based therapies. Cells 2023;12:1905.

35. Esquivel D, Mishra R, Srivastava A. Potential use of stem cell therapies for treating osteoarthritis and rheumatoid arthritis. Ann Transl Med 2024;12:72.

36. Feng Z, Yang Y, Liu X-Z et al. Application of cell therapy in rheumatoid arthritis: Focusing on the immunomodulatory strategies of mesenchymal stem cells. Internat Immunopharm 2025;147:114017.

37. Ding Q, Hu W, Wang R et al. Signaling pathways in rheumatoid arthritis: implications for targeted therapy. Signal Transduc Target Ther 2023;8:68.

38. Zhang C, You Z, pan Z et al. Integrated analysis of the relationship between metabolic pathways and immune infiltration in rheumatoid arthritis. Front Immunol 2025;16:1679356.

39. Nieminen P, Hamalainen W, Savinainen J et al. Metabolomics of synovial fluid and infrapatellar fat pad in patients with osteoarthritis or rheumatoid arthritis. Inflammation 2022;45:1101–1117.

40. Cheon H, Yu S-J, Yoo, DH et al. Increased expression of pro-inflammatory cytokines and metalloproteinase-1 by TGF-β1 in synovial fibroblasts from rheumatoid arthritis and normal individuals. Clin Exp Immunol 2002;127:547–552.

41. Sakuma M, Hatsushika K, Koyama K et al. TGF-β type I receptor kinase inhibitor down-regulates rheumatoid synoviocytes and prevents the arthritis induced by type II collagen antibody. Internat Immunol 2006;19:117–126.

42. Gonzalo-Gil E, Criado G, Santiago B et al. Transforming growth factor (TGF)-β signalling is increased in rheumatoid synovium but TGF-β blockade does not modify experimental arthritis. Clin Exp Immunol 2013;174:245–255.

43. Wilson TM, Bolt M, Stahly A et al. Transforming growth factor-beta is increased in sputum from individuals with rheumatoid arthritis-associated pulmonary fibrosis. Rheumatol 2025;

44. Bhamidipati K, McIntyre ABR, Kazerounian S et al. Spatial patterning of fibroblast TGFβ signaling underlies treatment resistance in rheumatoid arthritis. Nat Immunol 2025;27:556–571.

45. Thapa N, Lee B-H, Kim I-S. TGFBIp/βig-h3 protein: A versatile matrix molecule induced by TGF-β. Int J Biochem Cell Biol 2007:39:2183–2194.

46. Nam EJ, Sa KH, You DW, et al. Up-regulated transforming growth factor β-inducible gene h3 in rheumatoid arthritis mediates adhesion and migration of synoviocytes through αvβ3 integrin. Arthritis Rheum 2006;54:2734–2744.

47. Murphy-Ullrich JE, Suto MJ. Thrombospondin-1 regulation of latent TGF-β activation: A therapeutic target for fibrotic disease. Matrix Biol 2018;68:28–43.

48. Murphy-Ullrich JE. Thrombospondin 1 and its diverse roles as a regulator of extracellular matrix in fibrotic disease. J Histochem Cytochem 2019;67:683–699.

49. Rico MC, Manns JM, Driban JB et al. Thrombospondin-1 and transforming growth factor beta are pro-inflammatory molecules in rheumatoid arthritis. Trans Res 2008;152:85–98.

50. Rico MC, Rough JJ, Del Carpio-Cano FE et al. The axis of thrombospondin-1, transforming growth factor beta and connective tissue growth factor: an emerging therapeutic target in rheumatoid arthritis. Curr Vasc Pharmacol 2010;8:338–343.

51. Ohyama K, Kawakami A, Tamai M et al. Serum immune complex containing thrombospondin-1: a novel biomarker for early rheumatoid arthritis. Ann Rheum Dis 2012;71:1916–1917.

52. Suzuki T, Iwamoto N, Yamasaki S et al. Upregulation of thrombospondin 1 expression in synovial tissues and plasma of rheumatoid arthritis: role of transforming growth factor-β1 toward fibroblast-like synovial cells. J rheumatol 2015;42”943–947.

53. Kaur S, Roberts DD. Emerging functions of thrombospondin-1 in immunity. Sem Cell Devel Biol 2024;155:22–31.

54. Hu J, Wang X, Ge C et al. TSP-1-CD47-integrin α4β1 axis drives T cell infiltration and synovial inflammation in rheumatoid arthritis. Front Immunol 2025;16:1524304.

55. Wilson PG, Thompson JC, Webb NR. Serum amyloid A, but not C-reactive protein, stimulates vascular proteoglycan synthesis in a pro-atherogenic manner. Am J Pathol 2008;173:1902–1910.

56. Liu X, Song Y, Hu S et al. Serum amyloid A contributes to radiation-induced lung injury by activating macrophages through FPR2/Rac1/NF-κB pathway. Int J Biol Sci 2024;20:4951–4956.

57. Zhou J, Dai Y, Lin Y, Chen K. Association between serum amyloid A and rheumatoid arthritis: A systematic review and meta-analysis. Sem Arth Rheum 2022;52:151943.

58. Leung EL, Lai H-L, Li R-Z et al. Roles of serum amyloid A 1 protein isoforms in rheumatoid arthritis. Engineering 2022;10:174–182.

59. Zinellu A, Mangoni AA. The potential role of serum amyloid A as biomarker of rheumatic diseases: a systematic review and meta-analysis. Clin Exper Med 2024;24:141.

60. Horiguchi M, Ota M, Rifkin DB. Matrix control of transforming growth factor-β function. J Biochem 2012;152:321–329.

61. van Beers JJBC, Willemze A, Stammen-Vogelzangs J et al. Anti-citrullinated fibronectin antibodies in rheumatoid arthritis are associated with human leukocyte antigen-*DRB1* shared epitope alleles. Arthritis Res Ther 2012;14:R35.

62. Aarts J, van Camm A, Chen X et al. Local inhibition of TGF-β1 signaling improves Th17/Treg balance but not joint pathology during experimental arthritis. Sci Reports 2022;32:3182.

63. Jiang X, Jia Z, Yang B. Metabolomic analysis suggests thiamine monophosphate as a potential marker for mesenchymal stem cell transplantation outcomes in patients with SLE. Lupus Sci Med 2025;12:e001197.

